# Impact of Radiation on Cardiovascular Outcomes in Patients with Resectable Esophageal Cancer

**DOI:** 10.1101/19010835

**Authors:** Reith R Sarkar, Ahmadreza Hatamipour, Neil Panjwani, Patrick T Courtney, Daniel R Cherry, Mia A Salans, Anthony T Yip, Brent S Rose, Daniel R Simpson, Matthew P Banegas, James D Murphy

**Affiliations:** University of California San Diego School of Medicine, La Jolla, CA; University of California San Diego Department of Radiation Medicine and Applied Sciences, La Jolla, CA; Department of Radiation Oncology, Stanford University, Stanford, CA; Center for Health Research, Kaiser Permanente Northwest, Portland, OR

**Keywords:** Esophageal Cancer, Radiotherapy, Radiotherapy Adverse Effects, Cardiotoxicity, Cardiovascular Diseases

## Abstract

**Purpose:** Preoperative radiation therapy improves outcomes for operable esophageal cancer patients, though the proximity of the heart to the esophagus puts patients at risk of radiation-induced cardiovascular disease. This study characterizes the impact of radiation therapy and different radiation techniques on cardiovascular morbidity among a large cohort of esophageal cancer patients.

**Methods:** We identified 1,125 Medicare beneficiaries diagnosed between 2000 and 2011 with esophageal cancer who received surgery alone, or surgery preceded by either preoperative chemotherapy or preoperative chemoradiation. We used Medicare claims to identify severe adverse cardiovascular events in the perioperative and postoperative periods. Multivariable logistic regression and Fine-Gray models were used to determine the effect of pre-surgery treatment on the risk of perioperative and postoperative cardiovascular disease.

**Results:** Preoperative chemotherapy or preoperative chemoradiation did not significantly increase the risk of perioperative cardiovascular complications compared to surgery alone. Patients treated with preoperative chemoradiation had a 36% increased risk of having a postoperative cardiovascular event compared to patients treated with surgery alone (subdistribution hazard ratio [SDHR] 1.36; p=0.035). There was no significant increase in cardiovascular events among patients treated with preoperative chemotherapy (SDHR 1.18; p=0.40). Among patients treated with preoperative chemoradiation, those receiving intensity modulated radiotherapy (IMRT) had a 68% decreased risk of having a cardiovascular event compared to patients receiving conventional radiation (SDHR 0.32; p=0.007).

**Conclusions:** This study demonstrates an increased risk of cardiovascular complications among operative esophageal cancer patients treated with preoperative chemoradiation, though these risks might be reduced with more cardioprotective radiation techniques such as IMRT.

## Introduction

Radiation therapy represents a central component in the treatment of operable esophageal cancer^1^. Among patients with localized or local-regional disease, clinical trials demonstrate that the use of preoperative radiation delivered concurrently with chemotherapy followed by surgery confers a survival benefit compared with surgery alone^2,3^. The CROSS study in particular found a median survival of 49 months for patients treated with preoperative chemoradiation followed by surgery compared to 24 months for patients treated with surgery alone^3^.

The location of the esophagus with respect to the heart puts the heart at risk of receiving incidental radiation during treatment for esophageal cancer. With other cancer types, including breast cancer, lung cancer, and Hodgkin’s lymphoma, large clinical studies demonstrate a clear link between thoracic radiation and adverse cardiovascular outcomes^4-6^. Multiple small institutional studies have demonstrated a potential link between radiation and cardiovascular disease in esophageal cancer patients treated with radiotherapy^7-11^. Additionally, a study evaluating patients within the Surveillance Epidemiology and End Results (SEER) cancer registry found increased cardiovascular mortality among esophageal cancer patients receiving radiation^12^. While these older studies demonstrate increased risks associated with radiation, the delivery of radiation in esophageal cancer has evolved over the past several decades^13^. Modern conformal treatment techniques, such as intensity modulated radiotherapy (IMRT), offer the ability to decrease radiation doses to the heart^14^. The purpose of this large population-based study was to characterize adverse cardiovascular events associated with radiation among operable patients with esophageal cancer. Furthermore, this study will analyze whether IMRT mitigates risks of radiation-induced heart disease.

## Methods and Materials

### Data Source

We identified esophageal cancer patients from the SEER-Medicare linked database. The National Cancer Institute oversees the SEER program which collects data on incident cancers diagnosed across the United States covering approximately 28% of the US population. Medicare provides federally funded health insurance for people over the age of 65. The SEER-Medicare linkage provides Medicare claims data for Medicare beneficiaries within the SEER database. Medicare claims indirectly capture information about cancer treatments and long-term risks of adverse events associated with treatment. The Institutional Review Board at the University of California San Diego found this study to be exempt from review.

### Study Population

Our initial query of the SEER-Medicare database identified 10,353 patients with histologically confirmed non-metastatic esophageal cancer diagnosed between 2000 and 2011. We included only patients with invasive adenocarcinoma or squamous cell carcinoma histology. To help maintain focus on adverse outcomes associated with esophageal cancer, we excluded subjects with a diagnosis of more than one cancer. Patients were required to have continuous Medicare claims data from 1 year prior to diagnosis (to calculate pre-existing comorbidity) through death or the end of follow-up. This required patients to have continuous Medicare Part A and B coverage over this period, and we excluded subjects with Part C coverage because managed care organizations do not consistently submit detailed claims data. Additional selection criteria are described below, and the final study cohort included 1,125 subjects (see **Figure 1** for selection schema).

**Figure 1.**
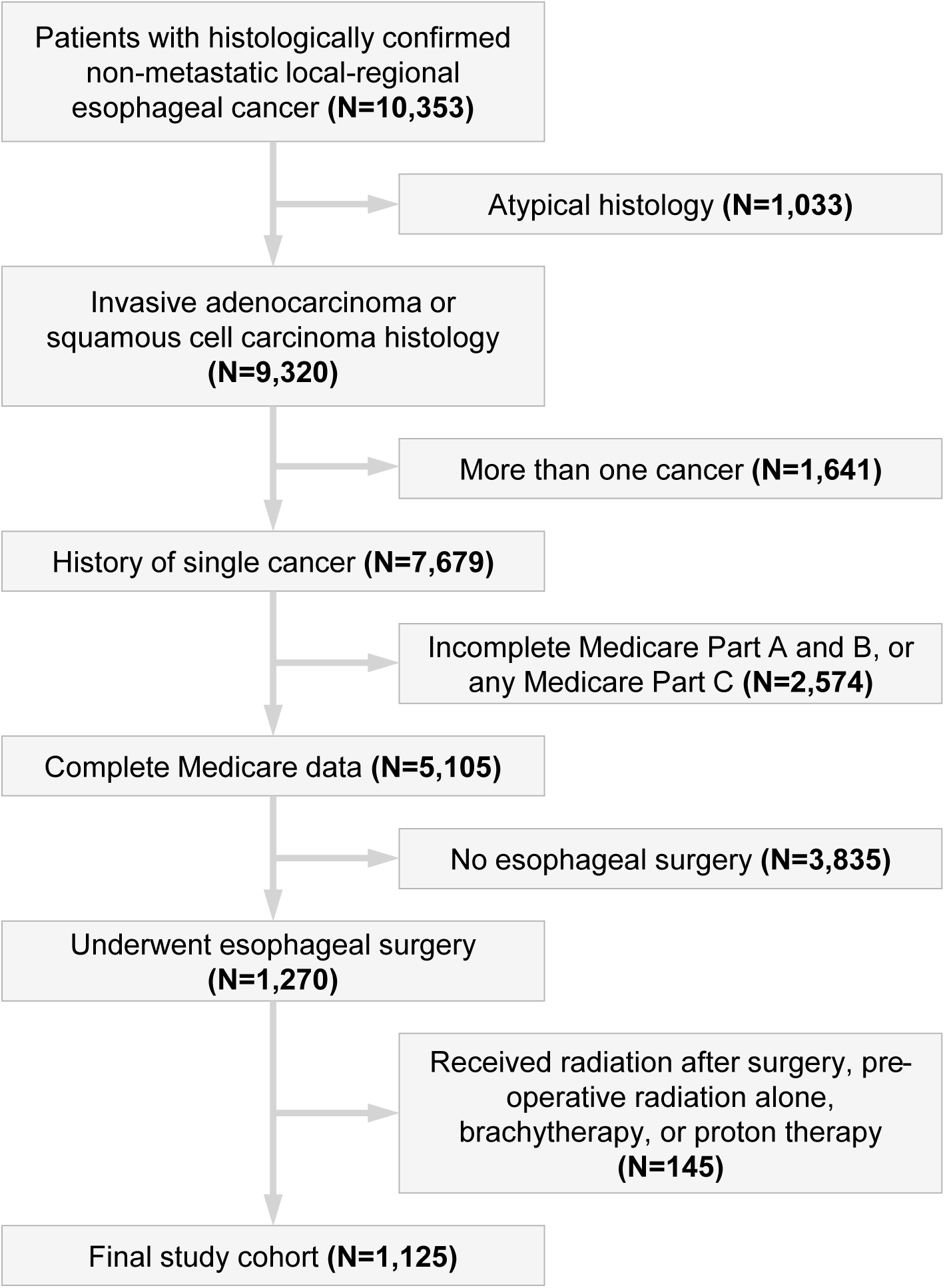
Patient selection.

### Study Covariates

Patient demographic and tumor characteristics including histology, stage, and year of diagnosis were extracted from SEER. We used the Deyo adaptation of the Charlson comorbidity index to calculate pre-existing comorbidity from inpatient and outpatient Medicare claims during the year prior to diagnosis^15^. This study separately incorporated pre-existing cardiovascular disease as a study variable (described below), therefore we modified the Charlson score by removing cardiovascular disease to avoid double counting. We identified chemotherapy, radiation, and esophageal surgery using previously established methods^16-18^ that rely on Medicare claims and International Classification of Diseases 9^th^ edition (ICD-9) Procedure and Diagnosis codes, as well as Healthcare Common Procedure Coding System (HCPCS) codes (see **Supplemental Table 1** for codes). We identified individual fractions of radiation from radiation treatment claims, and considered only definitive courses of radiation defined as 20 or more fractions of radiation. We assumed that a break between fractions of radiation of more than 30 days indicated a separate (and subsequent) course of radiation. We identified patients who received IMRT through radiation planning and treatment claims specific to IMRT. We defined concurrent chemoradiation as any administration of chemotherapy during or within a 2 week window surrounding the course of radiation. We excluded the few patients who received brachytherapy, or proton therapy. Additionally, we excluded the small number of patients who received preoperative radiation alone without concurrent chemotherapy (insufficient numbers for analysis), as well as patients receiving postoperative radiotherapy (see **Figure 1**).

### Cardiovascular events

The primary study endpoint included clinically relevant cardiovascular events potentially associated with radiation ascertained with ICD-9 and HCPCS codes (**Supplemental Table 1**). This composite endpoint included hospitalization associated with any of the following: acute myocardial infarction, percutaneous coronary intervention (PCI), coronary artery bypass grafting (CABG), valve surgery, pericardial disease, or congestive heart failure or cardiomyopathy. We classified cardiovascular endpoints as preexisting, perioperative, and postoperative based on timing with respect to cancer diagnosis and surgery. *Preexisting events* occurred during the year prior to cancer diagnosis. *Perioperative events* occurred between the date of esophageal surgery and 30 days after discharge from the hospital. *Postoperative events* occurred more than 30 days after discharge.

### Statistical analysis

We characterized the study cohort with median follow-up time, calculated from the date of surgery through death or last follow-up, and the median and 5-year survival, estimated with Kaplan-Meier analyses. We categorized patients into three groups based on the type of preoperative treatment received: preoperative chemotherapy alone, preoperative chemoradiation, or no preoperative treatment. Among these three treatment groups we assessed for differences in baseline patient and tumor characteristics with Fisher’s exact tests. We used multivariable models to determine whether the risks of perioperative and postoperative cardiovascular events varied by preoperative treatment. Multivariable models incorporated potential confounding factors including patient age, race, sex, marital status, year of diagnosis, tumor histology, stage (localized vs. regional), Charlson comorbidity score, preexisting cardiovascular disease, and the receipt of adjuvant chemotherapy. The impact of preoperative treatment on perioperative cardiovascular events was assessed with a multivariable logistic regression. The impact of preoperative treatment on postoperative cardiovascular events was assessed with a multivariable Fine-Gray regression considering death as a competing event, censoring at last follow-up. To determine whether the risk of postoperative cardiovascular events varied by patient characteristics, we performed an additional analysis introducing interaction terms with treatment and select variables into the multivariable model. A significant interaction would indicate that the impact of treatment on the risk of cardiovascular disease varied by that patient characteristic. The 5-year incidence of cardiovascular events were assessed with cumulative incidence analyses. Due to the small number of cardiac deaths (N=58) we did not assess cardiovascular mortality in this analysis. Statistical analyses were conducted with SAS version 9.4 (SAS Institute Inc., Cary, NC) with p-values < 0.05 considered significant.

## Results

Among the 1,125 esophageal cancer patients in this study the median follow-up time was 1.5 years, and for the patients alive at the end of the study (N=355; 32%), the median follow-up time was 5.3 years. The median survival for the entire cohort measured from the date of surgery was 1.9 years, and the 5-year survival was 33%. When considering preoperative treatment we found that 613 patients (54%) received surgery alone, 137 (12%) received preoperative chemotherapy, and 375 (33%) received preoperative chemoradiation. **Table 1** demonstrates patient characteristics stratified by treatment group. Compared to patients receiving surgery alone, the patients receiving chemoradiation were younger, more likely married, diagnosed at a later time period, were more likely to have regional disease on presentation, and were more likely to have received chemotherapy after surgery.

**Table 1.** Patient characteristics.

| Characteristic | N | Treatment group – N (%) |  |  | p-value |
| --- | --- | --- | --- | --- | --- |
|  |  | Surgery alone | Pre-operative chemotherapy | Pre-operative chemoradiation |  |
| Total | 1,125 | 613 | 137 | 375 |  |
| Age |  |  |  |  | <0.0001 |
| 65-74 | 750 | 360 (58.7) | 92 (67.2) | 298 (79.5) |  |
| 75+ | 375 | 253 (41.3) | 45 (32.9) | 77 (20.5) |  |
| Race |  |  |  |  | 0.46 |
| White | 1,034 | 562 (91.7) | 123 (89.8) | 349 (93.1) |  |
| Other | 91 | 51 (8.3) | 14 (10.2) | 26 (6.9) |  |
| Marital status |  |  |  |  | 0.0005 |
| Married | 773 | 392 (64) | 98 (71.5) | 283 (75.5) |  |
| Other | 352 | 221 (36.1) | 39 (28.5) | 92 (24.5) |  |
| Sex |  |  |  |  | 0.37 |
| Male | 868 | 463 (75.5) | 108 (78.8) | 297 (79.2) |  |
| Female | 257 | 150 (24.5) | 29 (21.2) | 78 (20.8) |  |
| Year of diagnosis |  |  |  |  | 0.0002 |
| 2000-2003 | 365 | 225 (36.7) | 50 (36.5) | 90 (24) |  |
| 2004-2007 | 402 | 216 (35.2) | 46 (33.6) | 140 (37.3) |  |
| 2008-2011 | 358 | 172 (28.1) | 41 (29.9) | 145 (38.7) |  |
| Histology |  |  |  |  | 0.99 |
| Squamous cell carcinoma | 283 | 153 (25) | 35 (25.6) | 95 (25.3) |  |
| Adenocarcinoma | 842 | 460 (75) | 102 (74.5) | 280 (74.7) |  |
| Stage |  |  |  |  | <0.0001 |
| Localized | 482 | 325 (53) | 50 (36.5) | 107 (28.5) |  |
| Regional | 643 | 288 (47) | 87 (63.5) | 268 (71.5) |  |
| Charlson comorbidity index |  |  |  |  | 0.98 |
| 0 | 680 | 370 (60.4) | 84 (61.3) | 226 (60.3) |  |
| 1+ | 445 | 243 (39.6) | 53 (38.7) | 149 (39.7) |  |
| Pre-existing cardiovascular disease |  |  |  |  | 0.24 |
| No | 1,070 | 577 (94.1) | >126 (92.0) | 362 (96.5) |  |
| Yes | 55 | 36 (5.9) | <11 (9.0) | 13 (3.5) |  |
| Chemotherapy after surgery |  |  |  |  | <0.0001 |
| No | 891 | 545 (88.9) | 91 (66.4) | 255 (68) |  |
| Yes | 234 | 68 (11.1) | 46 (33.6) | 120 (32) |  |

In the perioperative period, 302 patients (27%) experienced a cardiovascular event, with the majority of these classified as congestive heart failure (N=259; 23%), or acute myocardial infarction (N=61; 5.4%). On multivariable analysis, we found that the risk of perioperative cardiovascular events did not correlate with the type of preoperative treatment. Compared to the surgery alone group, there was no increased risk of developing a perioperative cardiovascular event among those receiving preoperative chemotherapy (odds ratio (OR) 0.83, 95% confidence interval (CI) 0.54-1.27; p=0.50), or among those receiving preoperative chemoradiation (OR 0.92, 95% CI 0.67-1.26; p=0.97).

During the postoperative period, the 5-year cumulative incidence of cardiovascular events among all patients was 25.7% (**Figure 2**), with most of the events consisting of congestive heart failure, pericardial disease, or myocardial infarction (**Table 2**). Multivariate analysis found that patients treated with preoperative chemoradiation had a 36% increased risk of developing a cardiovascular event compared to those undergoing surgery alone (sub-distribution hazard ratio [SDHR] 1.36, 95% CI 1.02-1.80; p=0.035). We did not find a significant increase in postoperative cardiovascular events risk among those receiving preoperative chemotherapy (SDHR 1.18, 95% CI 0.80-1.75; p=0.40). The impact of preoperative chemoradiotherapy on postoperative cardiovascular outcomes did not differ according to patient age, race, sex, marital status, year of diagnosis, tumor histology, stage (localized vs. regional), Charlson comorbidity score, or preexisting cardiovascular disease (all interaction p-values >0.05).

**Table 2.**
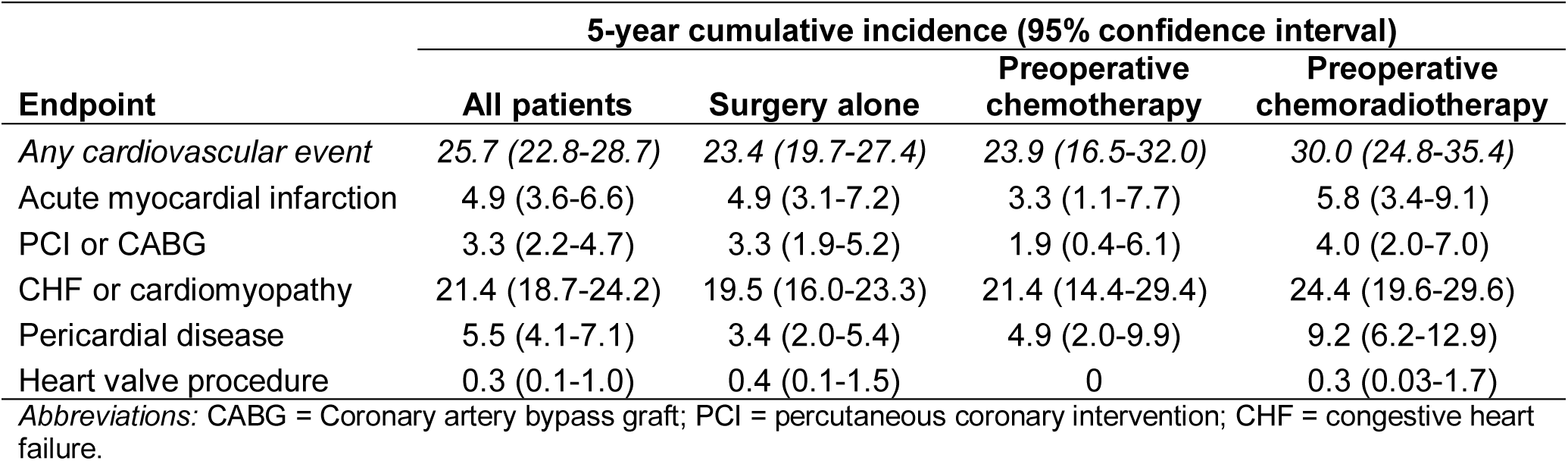
Postoperative cardiovascular events.

| **Endpoint** | **5-year cumulative incidence (95% confidence interval)** |  |  |  |
| --- | --- | --- | --- | --- |
|  | **All patients** | **Surgery alone** | **Preoperative chemotherapy** | **Preoperative chemoradiotherapy** |
| *Any cardiovascular event* | 25.7 (22.8-28.7) | 23.4 (19.7-27.4) | 23.9 (16.5-32.0) | 30.0 (24.8-35.4) |
| Acute myocardial infarction | 4.9 (3.6-6.6) | 4.9 (3.1-7.2) | 3.3 (1.1-7.7) | 5.8 (3.4-9.1) |
| PCI or CABG | 3.3 (2.2-4.7) | 3.3 (1.9-5.2) | 1.9 (0.4-6.1) | 4.0 (2.0-7.0) |
| CHF or cardiomyopathy | 21.4 (18.7-24.2) | 19.5 (16.0-23.3) | 21.4 (14.4-29.4) | 24.4 (19.6-29.6) |
| Pericardial disease | 5.5 (4.1-7.1) | 3.4 (2.0-5.4) | 4.9 (2.0-9.9) | 9.2 (6.2-12.9) |
| Heart valve procedure | 0.3 (0.1-1.0) | 0.4 (0.1-1.5) | 0 | 0.3 (0.03-1.7) |

*Abbreviations:* CABG = Coronary artery bypass graft; PCI = percutaneous coronary intervention; CHF = congestive heart failure.

**Figure 2.**
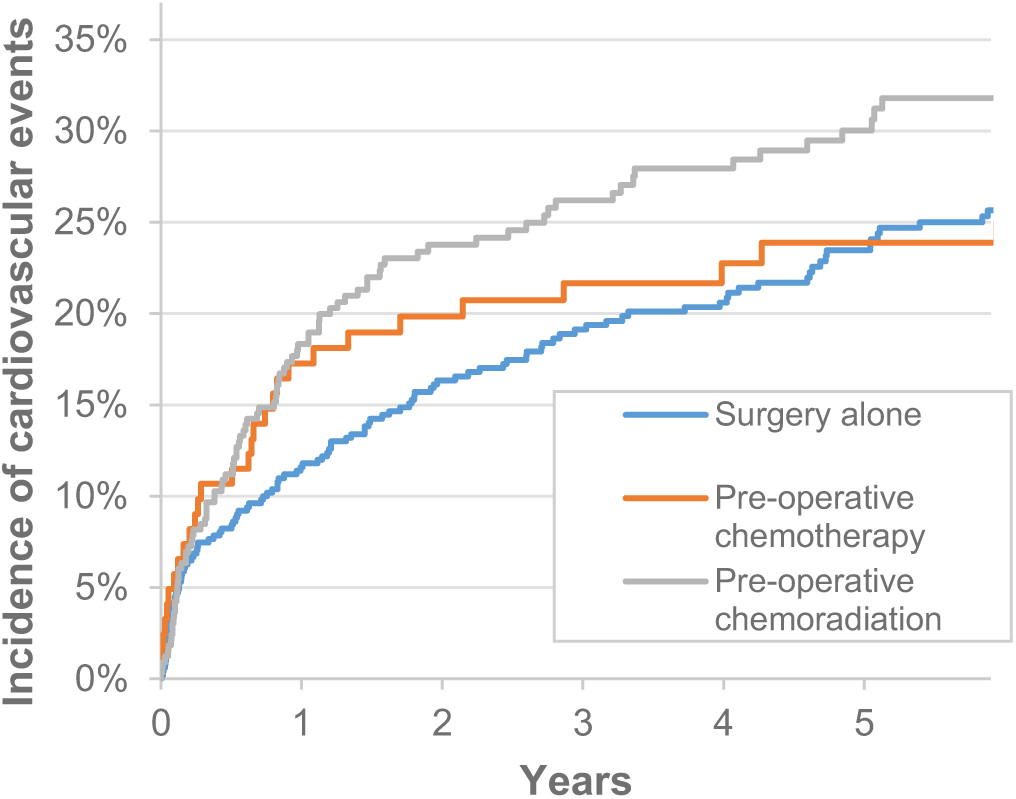
Post-operative cardiovascular events.

Among the cohort of patients receiving chemoradiotherapy, 71 (19%) received IMRT. The 5-year cumulative incidence of postoperative cardiovascular events was 12.1% (95% CI 5.1-22.2%) for patients receiving IMRT compared to 33.2% (95% CI 27.3-39.2%) for patients receiving conventional radiation (**Figure 3**). On multivariable analysis, patients receiving IMRT had a 68% decreased risk of having a postoperative cardiovascular event compared to patients receiving conventional radiation (SDHR 0.32, 95% CI 0.14-0.73; p=0.007).

**Figure 3.**
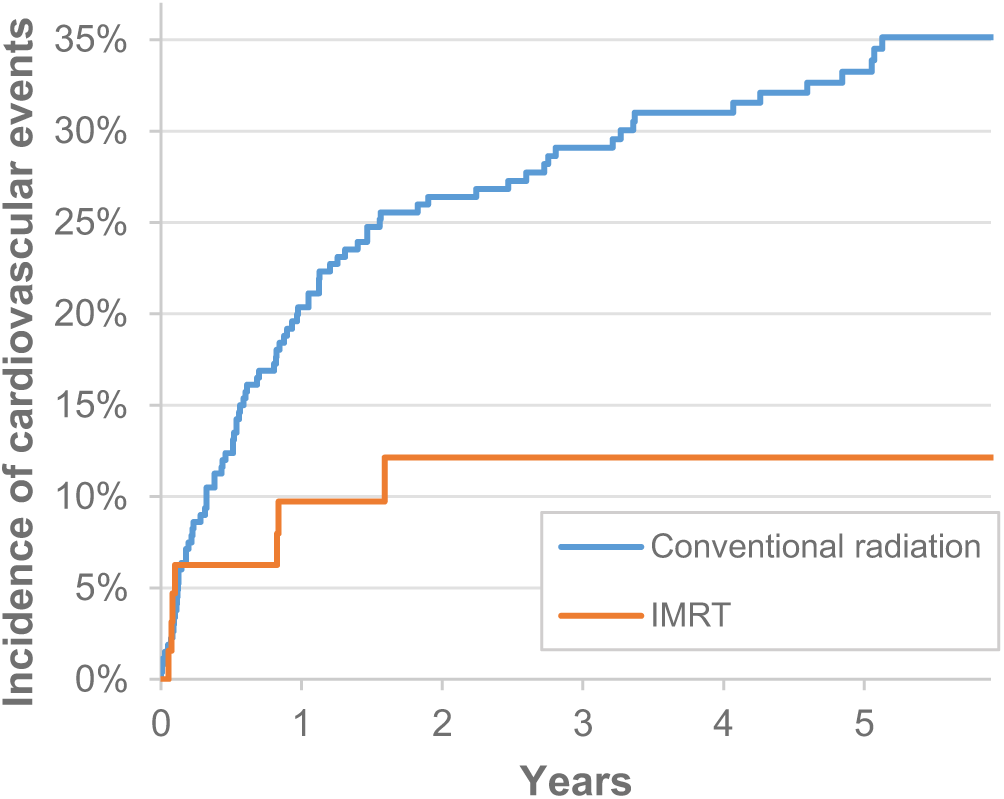
Impact of radiation modality on postoperative cardiovascular events.

## Discussion

This study demonstrates a link between the receipt of radiation and long-term postoperative cardiovascular outcomes among operable esophageal cancer patients receiving curative intent treatment. Our key finding of a 36% increased relative risk of adverse, clinically relevant cardiovascular events supports the existing literature evaluating the risk of radiation-induced cardiotoxicity in esophageal cancer. A series of single institution studies with sample sizes ranging from 15 to 123 have described a similar association between radiation in esophageal cancer and a variety of cardiovascular endpoints. This includes studies identifying a link between radiation and increased myocardial perfusion defects^7^, decreased ejection fraction^8,9^, and clinically significant cardiac toxicity^10,11^. Similarly, a study evaluating patients within SEER found a 2.8% absolute increased risk of death from cardiovascular disease among esophageal cancer patients treated with radiation^12^. Clinical trial data support a cancer-specific survival advantage associated with preoperative radiation in esophageal cancer^3,19^, though our study in conjunction with existing research highlights the potential for increased cardiovascular morbidity and mortality.

The timing of cardiovascular events after radiation deserves further discussion. Imaging research demonstrates that myocardial perfusion defects can occur as early as 6 months after treatment^20-22^. Furthermore, research in breast cancer demonstrates that the long-term cardiovascular effects of radiation stretch over a patient’s lifetime^4,23^. The probability of long-term survival with esophageal cancer is lower than breast cancer, though given the trend towards improved survival over time with esophageal cancer^24^, one could argue that the clinical importance of radiation-induced cardiovascular disease among esophageal cancer survivors will increase in the future.

Another key finding of this study relates to the observed reduction in postoperative cardiovascular events among patients receiving IMRT. Research evaluating radiation dosimetry demonstrates that the more conformal radiation dose distributions with IMRT have the potential to reduce radiation doses to the heart compared to conventional forms of radiation^14,25-28^. Similarly, other retrospective analyses have found potential reductions in cardiovascular toxicity associated with IMRT compared to older radiation techniques^29,30^. Furthermore, a cancer registry study found that IMRT was associated with a decreased risk of cardiac mortality compared to conventional radiation^31^. The potential benefits of more conformal radiation modalities raise the question about the possible utility of proton therapy in esophageal cancer, which can further reduce radiation doses to the heart^32^. A recent early report of a phase II randomized trial comparing proton therapy to IMRT demonstrated reduced toxicity among the proton therapy patients, though did not specifically report on cardiovascular toxicity^33^. An ongoing phase III randomized trial with a co-primary endpoint of overall survival and adverse cardiopulmonary events will help define the potential benefit of proton therapy in esophageal cancer (NCT03801876).

An important consideration of this study relates to the potential contribution of chemotherapy on the risks of adverse cardiovascular events. Combined chemotherapy and radiation represents the standard approach in the definitive management of non-metastatic esophageal cancer^1^. The typical chemotherapy doublets used concurrently with radiation in esophageal cancer include 5-fluorouracil and cisplatin, or carboplatin and paclitaxel^1^. 5-fluorouracil can cause coronary spasms or ischemia, and paclitaxel can cause bradycardia^34^. Furthermore, all of these chemotherapy agents have the capacity for radiosensitization^35^, which could influence the adverse impact of radiation on the heart. Whether different concurrent chemotherapy agents delivered with radiation lead to different risks of cardiotoxicity remains an important but unanswered question.

This study has limitations worth mentioning. The SEER-Medicare linked database lacks specific details of radiation including the radiation dose and target, both of which could impact the amount and anatomic distribution of radiation received by the heart, which could influence the patient-specific risks of cardiovascular disease. Furthermore, this study lacked data on important cardiovascular risk factors including smoking, obesity, exercise, and family history, any of which could mitigate or moderate the risk of cardiovascular morbidity and mortality. This study included Medicare beneficiaries over the age of 65, therefore the findings in this study may not generalize to a younger population. Finally, with the increased knowledge and awareness surrounding radiation-induced heart disease, one could hypothesize that radiation oncologists treating patients today might prioritize cardiac sparing in their radiation plans, which could decrease the risks of radiation-induced heart disease for patients treated in current practice.

Despite these limitations, this population-based study demonstrates that preoperative chemoradiation increases the risk of cardiovascular complications among esophageal cancer patients treated with curative intent. Furthermore, this study demonstrates the capacity of more advanced radiation techniques such as IMRT to reduce these risks, which provides a rationale to use cardioprotective radiation techniques. While additional research incorporating radiation dose is needed to quantify patient-specific risks, the findings of this study can help patients and radiation oncologists better understand the risks of radiation in esophageal cancer.

## Data Availability

All data referred to in the manuscript is publicly available through the SEER-Medicare linked database.

